# Spatiotemporal dynamics of hippocampal-cortical networks underlying the unique phenomenological properties of trauma-related intrusive memories

**DOI:** 10.1101/2023.06.20.23291671

**Authors:** Kevin J. Clancy, Quentin Devignes, Boyu Ren, Yara Pollmann, Sienna R. Nielsen, Kristin Howell, Poornima Kumar, Emily L. Belleau, Isabelle M. Rosso

## Abstract

Trauma-related intrusive memories (TR-IMs) possess unique phenomenological properties that contribute to adverse post-traumatic outcomes, positioning them as critical intervention targets. However, transdiagnostic treatments for TR-IMs are scarce, as their underlying mechanisms have been investigated separate from their unique phenomenological properties. Extant models of more general episodic memory highlight dynamic hippocampal-cortical interactions that vary along the anterior-posterior axis of the hippocampus (HPC) to support different cognitive-affective and sensory-perceptual features of memory. Extending this work into the unique properties of TR-IMs, we conducted a study of eighty-four trauma-exposed adults who completed daily ecological momentary assessments of TR-IM properties followed by resting-state functional magnetic resonance imaging (rs-fMRI). Spatiotemporal dynamics of anterior and posterior hippocampal (a/pHPC)-cortical networks were assessed using co-activation pattern analysis to investigate their associations with different properties of TR-IMs. Emotional intensity of TR-IMs was inversely associated with the frequency and persistence of an aHPC-default mode network co-activation pattern. Conversely, sensory features of TR-IMs were associated with more frequent co-activation of the HPC with sensory cortices and the ventral attention network, and the reliving of TR-IMs in the “here-and-now” was associated with more persistent co-activation of the pHPC and the visual cortex. Notably, no associations were found between HPC-cortical network dynamics and conventional symptom measures, including TR-IM frequency or retrospective recall, underscoring the utility of ecological assessments of memory properties in identifying the neural substrates of memory properties. These findings provide novel mechanistic insights into the unique features of TR-IMs that are critical for the development of individualized, transdiagnostic treatments for this pervasive, difficult-to-treat symptom.

## INTRODUCTION

Intrusive memories (IMs) of a traumatic experience are common among trauma-exposed individuals and are predictors of the onset, maintenance, and severity of transdiagnostic post-traumatic sequalae [1–3]. As such, IMs are positioned as critical intervention targets for survivors of trauma [4, 5]. However, mechanism-based treatments for trauma-related (TR)-IMs are scarce, due in part to a lack of biological models that account for their unique phenomenological properties.

Transdiagnostically, TR-IMs are involuntary, spontaneous, and intrude on conscious thought. They are characterized by vivid sensory fragments of the trauma that emerge with deficient contextual details, such as place and time. These sensory-perceptual properties contribute to a sense of reliving in the “here-and-now” and distinguish IMs from other forms of episodic memory [6]. Additionally, IMs exhibit distinct cognitive-affective properties, such as significant emotional distress and “attentional hijacking” [7, 8]. These phenomenological properties have given rise to various conceptual models of TR-IMs have emerged from these phenomenological properties, especially within the context of posttraumatic stress disorder (PTSD). The “warning signal hypothesis” emphasizes the exaggerated affective valence of TR-IMs as a learned cue to acquired threat, such that emotional qualities of TR-IMs may serve to activate threat-orienting and -responding behaviors in response to a (mis)perceived threat cue [9, 10]. Another model, the “dual-representation theory”, expands more on the sensory-perceptual features of IMs, proposing two parallel, yet interacting, memory systems: 1) a higher-order contextual representation (C-rep) system that stores contextual details of the event, and 2) a lower-order sensory representation (S-rep) system that stores sensory-perceptual features [6, 11]. This theory suggests IMs lack the necessary C-reps to bind exaggerated S-reps in place and time, thereby contributing to their sensory-rich re-experiencing in the “here and now”. Overall, these models emphasize the critical role phenomenological properties play in understanding the underlying mechanistic processes of TR-IMs.

These conceptual models have inspired neurobiological accounts of TR-IMs that, while scarce, are grounded in decades of neurocognitive models of episodic memory [12]. The dual representation theory of TR-IMs positions a hyperactive sensory cortex and salience network (SN) as hubs of the exaggerated S-rep system. This proposition is supported by accruing evidence implicating the sensory cortex in trauma memory and intrusions [13–16] and the long-term storage of conditioned threat in anxiety [17, 18]. Conversely, dysfunction of the hippocampal complex is believed to underpin the deficient C-rep system, given the well-established role of the hippocamps (HPC) in the contextual binding of memory features [19, 20]. More broadly, episodic memory is associated with dynamic interactions between the HPC and large-scale networks distributed across the brain, such as the “cortical retrieval”, default mode (DMN), and dorsal attention networks (DAN), as well as sensory, posterior-medial, and anterior-temporal systems [21–24]. These distributed networks support different aspects of episodic memory [25], and their functional segregations are mirrored in their distinct connectivity and co-activation patterns with anterior and posterior segments of the HPC [24, 26–30]. Specifically, the posterior HPC (pHPC) has been linked to posterior-medial and sensory systems supporting the detailed sensory-perceptual, particularly visuospatial, properties of memory and associated mental imagery [31–33]. In contrast, the anterior HPC (aHPC) is predominantly connected to prefrontal and limbic structures supporting more complex cognitive-affective features, including emotion and schematic gist [34, 35]. Taken together, the dynamic interactions of these distributed systems with anterior-posterior divisions of the HPC are uniquely positioned to support the different cognitive-affective versus sensory-perceptual properties of TR-IMs.

To date, the unique properties of TR-IMs have been investigated largely independently of their neurobiological substrates. The majority of neuroimaging research on TR-IMs has focused on the presence, frequency, and intensity of (experimentally-induced) IMs, consistent with clinical assessments of the symptom, and implicate a diverse set of cortical regions linked to sensory, cognitive, and affective processes [15, 36–38]. Additionally, the HPC is often examined as a static and unitary structure in such research. Evidence is accruing for differential impacts of a/pHPC structure and function in PTSD-related symptomatology [39–42], reflecting the functional heterogeneity of hippocampal subregions that may contribute to the diverse presentations of intrusion symptoms. Moreover, static connectivity measures between the unitary HPC and cortical structures may fail to capture the known dynamic nature of interactions between HPC subregions and distributed cortical systems [43, 44]. The intrinsic dynamics of these HPC-cortical interactions is of particular relevance to clinical presentations, as IM’s emerge spontaneously and intermittently from a “resting state” and intrinsic neural activity can predict an individual’s response to stimuli or cues [45–48].

Therefore, the objective of this study was to identify spatiotemporal patterns of dynamic intrinsic HPC-cortical co-activation that are associated with the different phenomenological properties of TR-IMs. We combined daily ecological momentary assessments of TR-IM properties with functional imaging of resting-state dynamic a/pHPC-cortical networks in trauma-exposed adults to test the hypothesis that the dynamics of different aHPC- and pHPC-cortical co-activation patterns would be associated with different TR-IM properties. Specifically, we hypothesized that sensory-perceptual properties would be associated with co-activation of the pHPC with the sensory cortex and SN, while cognitive-affective features would be associated with covariance of the aHPC with higher-order systems that regulate attention, self-reference, and emotion.

## METHODS AND MATERIALS

### Participants

Ninety-nine (99) trauma-exposed adults completed daily ecological momentary assessments (EMAs) of TR-IMs followed by resting-state functional magnetic resonance imaging (rs-fMRI) as part of a larger study. Participants were recruited and enrolled via advertisements in the local community. Study procedures were approved by the Mass General Brigham Human Research Committee and all participants provided written informed consent at Visit 1. Participants were recruited based on exposure to a Criterion A traumatic event and the endorsement of at least two TR-IMs per week over the past month, as defined by the DSM-5 [49]. Additional inclusion and exclusion criteria are provided in the Supplement.

Participants completed 2 weeks of daily ecological momentary assessments (EMAs) of TR-IMs. After these 2 weeks, they returned for Visit 2 to complete a clinical interview, self-report questionnaires, and a 13-minute eyes-open rs-fMRI scan. Of the 99 participants who completed the protocol, 84 had usable fMRI data (excluded: excessive motion = 10, structural abnormalities in regions of interest = 2, poor structural-functional alignment = 1, falling asleep during scan = 2), resulting in our final sample of N = 84. Demographic and clinical details of the analyzed sample are provided in Table 1.

**Table 1.**
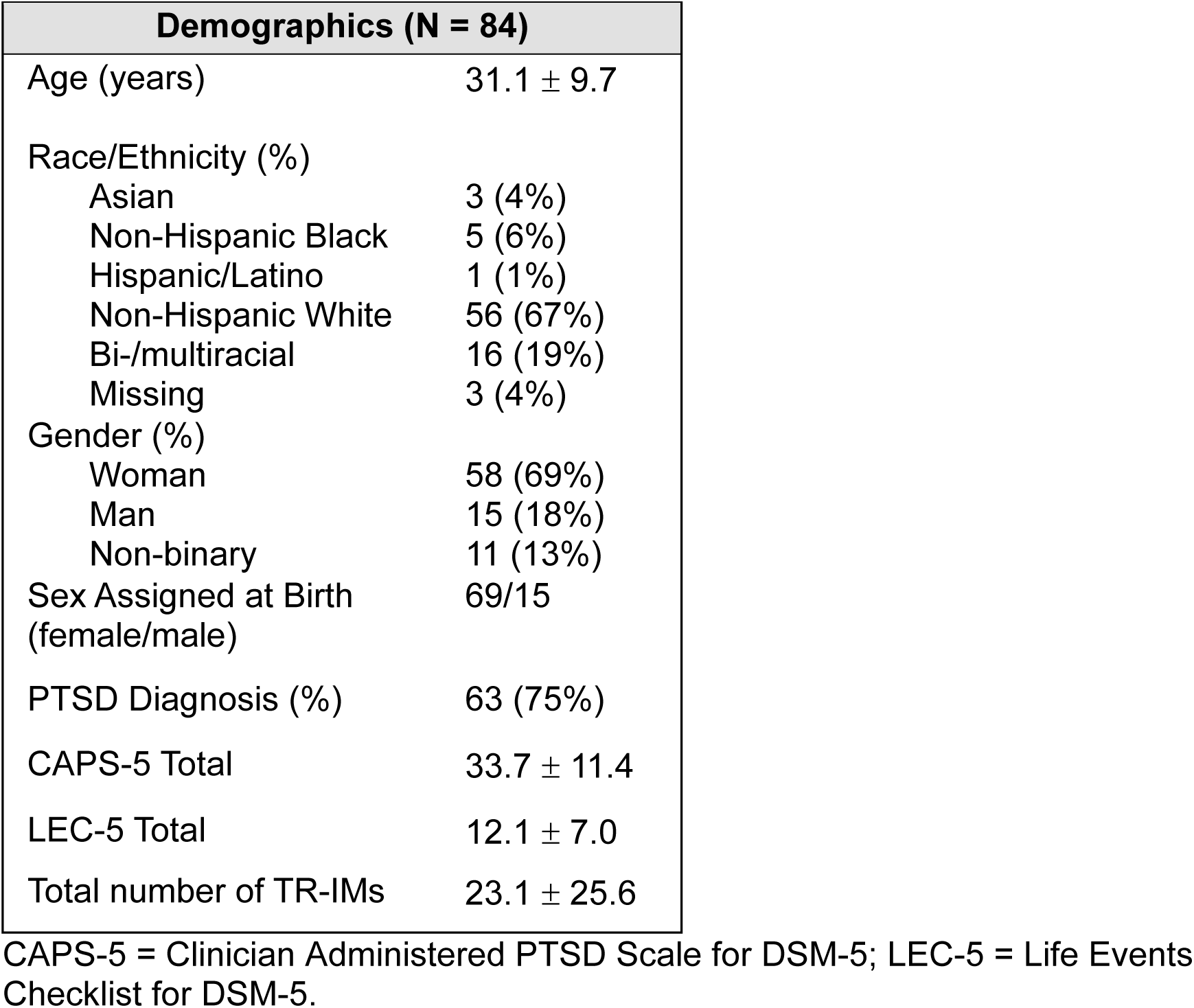
Demographic and clinical characteristics. Means ± standard deviations or N (%).

### Ecological momentary assessments

Participants completed daily EMAs of the phenomenological properties of TR-IMs for 2 weeks. The EMAs consisted of 3 daily surveys assessing TR-IMs for a maximum of 21 surveys per week. On a semi-random schedule, participants were prompted to complete these surveys on their smartphone using the MetricWire app. Surveys assessed for the presence of TR-IMs since the last survey, followed by 18 prompts about their phenomenological properties. Prompts about the properties were adapted from the Autobiographical Memory Questionnaire (AMQ) [50, 51] and were rated on a 0-4 Likert scale. Ratings were grouped into vividness, visual detail, reliving (here-and-now), emotional intensity, fragmentation, and intrusiveness, consistent with prior work [52].

### Interview and self-report measures

#### Clinician-Administered PTSD Scale for DSM-5 (CAPS-5)

The CAPS-5 [53], the gold-standard diagnostic interview for PTSD, was administered by doctoral-level clinicians during Visit 2. This interview consists of 30 items designed to assess the onset, duration, and impact of PTSD symptoms, yielding a determination of PTSD diagnosis and symptom severity, including specific symptom clusters.

#### Life Events Checklist (LEC-5)

The LEC-5 [54] is a 17-item assessment of potentially traumatic events used to determine which events a participant has experienced, witnessed, or learned about happening to a family member or close friend, reflecting a Criterion A trauma.

#### Autobiographical Memory Questionnaire (AMQ)

The AMQ [50, 51] is a 32-item questionnaire of autobiographical memory qualities, including categories of vividness, visual and auditory features, other sensory features, bodily sensations, language, emotions, perspective, nowness, fragmentation, and intrusiveness. Participants completed this survey during Visit 2, after completing the 2-week EMA-survey period, and retrospectively rated the qualities of their most intrusive and unwanted trauma-related memories on a 0-4 Likert scale.

### MRI data acquisition and preprocessing

Imaging was conducted at the McLean Hospital Imaging Center on a 3T Siemens Prisma scanner with a 64-channel head coil. Structural and functional images were acquired using the Human Connectome Project (HCP) Young Lifespan protocols [55], including a 13-minute eyes-open resting-state scan. Protocol details are documented in the Supplement. MRI data were preprocessed using fMRIPrep version 20.2.7 [56], including susceptibility distortion correction, co-registration, slice-time correction, normalization, motion correction, and spatial smoothing. Participants were excluded if their total mean framewise displacement (FD) across volumes exceeded 0.5 mm or greater than 20% of volumes exceeded FD = 0.5 mm (n = 10) [57]. Additional preprocessing of resting-state fMRI data was conducted using the CONN toolbox [58], including the regression of physiological noise from white matter and cerebrospinal fluid using the CompCor method [59], scrubbing of motion outliers (FD > 0.5 mm) [57], and high pass (0.01 Hz) filtering. Further preprocessing details are presented in the Supplement.

### Co-activation Pattern Analysis

Co-activation pattern (CAP) analysis [60] was used to compute spatiotemporal dynamics of anterior and posterior hippocampus (a/pHPC) networks. a/pHPC ROIs were defined based on prior work [39] using the SPM12 Anatomy Toolbox to maximize anterior-posterior segregation and minimize overlap with adjacent structures (i.e., amygdala). Additional a/pHPC ROI details are provided in the Supplement. Analyses were performed using functions derived from the TbCAP Toolbox [61]. A union seed-based approach was used to identify volumes that exceeded an activation threshold Z > 1 [60, 62, 63] of either the aHPC, pHPC, or both to ensure only volumes, or “states”, characterized by HPC activation were evaluated. Spatial patterns of co-active regions within selected volumes were then clustered into co-activation patterns (CAPs) using k-means clustering. K-means consensus clustering was performed to determine the optimal number of CAPs within the data [64, 65] and identified *k* = 4 as optimal. K-means clustering was run 100 times to avoid local minima and ensure stability of the 4 CAPs. Additional details on k-means clustering and consensus clustering are provided in the Supplement.

We computed the following CAP metrics within each participant: 1) *count*, reflecting the total supra-threshold volumes characterized by each CAP, and 2) *persistence*, reflecting the probability to remain in a given CAP across consecutive volumes. No effects were seen with total number of supra-threshold volumes across all CAPs (*p*’s > 0.07) and results were virtually identical when using fractional CAP count, which controls for individual differences in total number of supra-threshold volumes (Supplemental Results).

### Statistical Analyses

The cross-sectional associations between CAP metrics and TR-IM properties were evaluated using partial correlations of TR-IM property ratings averaged across the survey period, controlling for the effect of age and sex. Correction for multiple comparisons was performed for each CAP metric using false discovery rate (FDR) at two levels – across all tests performed (4 CAPs x 6 TR-IM properties = 24 tests; FDR_total_) and across CAPs within each TR-IM property (4 CAPs x 1 TR-IM property = 4; FDR_property_). Averaged TR-IM properties demonstrating a significant effect were then entered as dependent variables in separate multiple linear regression models with all CAPs as predictors to demonstrate a specificity of the association between individual TR-IM properties and CAPs.

To evaluate the robustness of the cross-sectional results, repeated measures over time of TR-IM properties demonstrating a significant cross-sectional association were analyzed by linear mixed models (LMM) with fixed effects for CAP metrics and a subject-specific random intercept to examine whether the same associations are present longitudinally. LMMs were used to account for the intra-subject correlations among the repeated measures. Two types of LMMs were considered: 1) univariate LMMs where CAP metrics were included individually and 2) multivariate LMMs where multiple CAP metrics are simultaneously included in the model. The analysis was performed in R version 4.2.2 (http://r-project.org/) using the lme4 package [66]. TR-IM properties and CAP metrics were mean-centered and scaled. The same FDR correction as in the cross-sectional analysis was applied to the results of univariate LMMs to adjust for multiple comparisons.

Finally, correlation analyses were performed between CAP metrics and conventional clinical symptom measures, including total number of TR-IMs (frequency), cross-sectional AMQ data of TR-IM properties at Visit 2 (retrospective recall), and CAPS-5 symptom severity, to demonstrate the utility of ecological assessments of TR-IMs.

## RESULTS

### CAP Characteristics

The CAPs consisted of coactivation of the HPC with activation of the DMN and deactivation of the SN/VAN and DAN (CAP1), activation (CAP2) and deactivation (CAP3) of the visual cortex (VC), and a pattern of SN, VC, and sensorimotor activation with DMN deactivation (CAP4; Figure 1A). CAP1 had more occurrences and persistence than all other CAPs (t’s > 3.82, p’s < 0.001), and CAP3 was more persistent than CAP4 (t = 2.90, p = 0.005; Figure 1B). There were no other differences in CAPs count or persistence.

**Figure 1.**
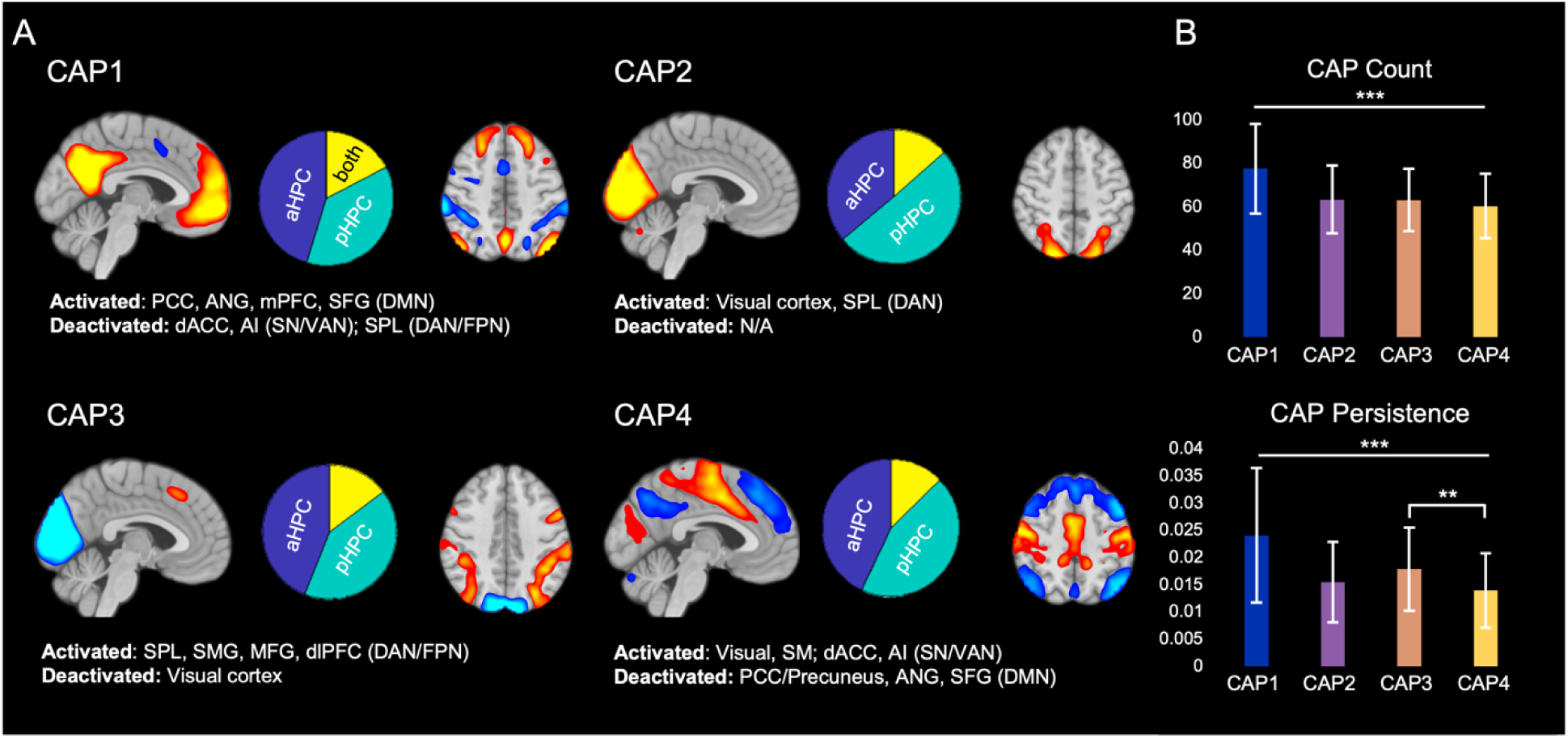
Representation of CAPs and their properties, including A) a summary of activated/deactivated regions and proportion of a/pHPC co-activation and B) the average count and persistence of each CAP across participants. PCC = posterior cingulate cortex, ANG = angular gyrus, mPFC = medial prefrontal cortex, SFG = superior frontal gyrus, DMN = default mode network, dACC = dorsal anterior cingulate cortex, AI = anterior insula, SN = salience network, VAN = ventral attention network, SPL = superior parietal lobule, DAN = dorsal attention network, FPN = frontoparietal network, SMG = supramarginal gyrus, MFG = middle frontal gyrus, dlPFC = dorsolateral prefrontal cortex, SM = sensorimotor cortex. ** *p* < 0.01, *** *p* < 0.001. Error bars reflect standard deviation.

Consistent with their divergent intrinsic connectivity networks, the aHPC and pHPC were differentially associated with the CAPs (Figure 1A): CAP1 was predominantly associated with activation of the aHPC [mean volume counts (fraction volume counts) ± S.D. = 35.5 (45%) ± 11.1 vs. pHPC: 28.1 (37%) ± 8.1; t = 6.27, p < 0.001], while CAP2 was dominated by pHPC activation [32.2 (50%) ± 7.8 vs. aHPC: 22.7 (36%) ± 7.5; t = 11.3, p < 0.001]. CAP3 and CAP4 were associated with equivalent a/pHPC activation (p’s > 0.10).

### CAP Count and TR-IM Properties

Visual properties were associated with more occurrences of CAP4 (HPC – VC/SM/dACC/AI; *r*_partial_ = 0.33, *p* = 0.002, FDR_total_ *p* < 0.05; Figure 2B), and emotional intensity was associated with fewer occurrences of CAP1 (aHPC-DMN; *r*_partial_ = -0.32, *p* = 0.003, FDR_total_ *p* < 0.05; Figure 2C). Individual linear regressions for each significant TR-IM property testing the specificity of its associations with individual CAPs revealed a unique association between visual properties and CAP4 occurrences (b = 0.026, *p* = 0.009) but did not demonstrate a specific association between emotional intensity and CAP1 (*p* = 0.668).

**Figure 2.**
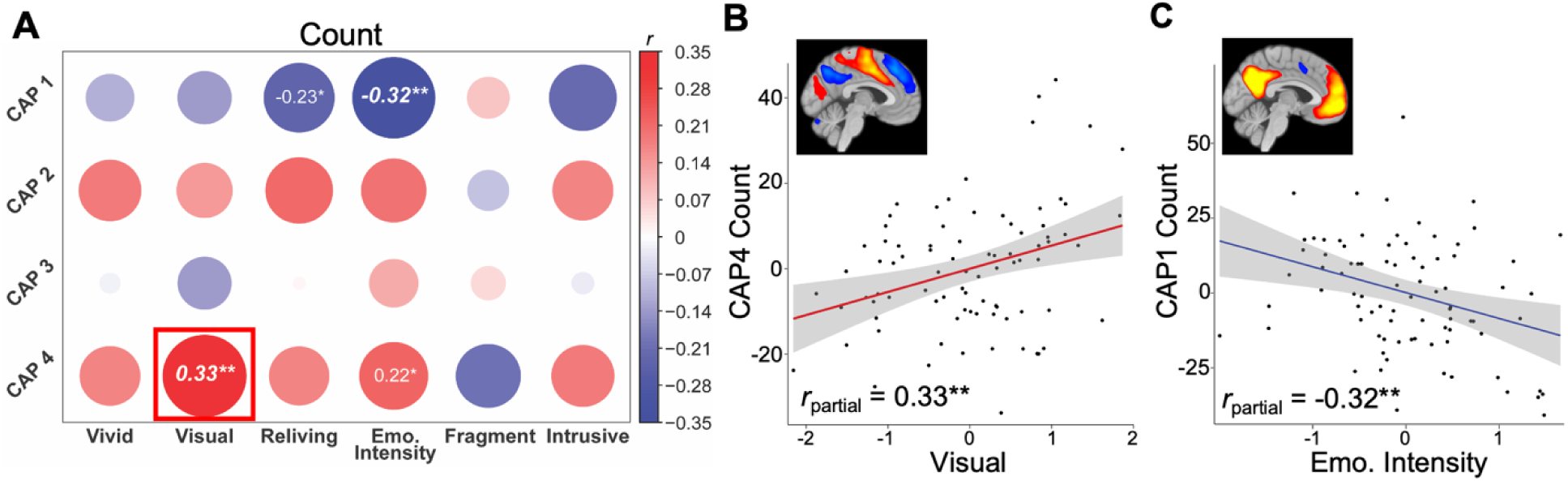
Associations between CAP Count and TR-IM properties. A) Partial correlations between count of all CAPs and TR-IM properties, controlling for age and sex, with the specific scatter plots of B) CAP4 and visual features and C) CAP1 and emotional intensity. Bold italics denote associations surviving correction for multiple comparisons. Boxes denote associations that were significant in multiple linear regression models, demonstrating specific association between that CAP and TR-IM property. * *p* < 0.05, ** *p* < 0.005.

Results from univariate LMMs confirmed these associations – visual properties were associated with more CAP4 occurrences (b = 0.28, *t* = 3.11, *p* = 0.003, FDR_total_ *p* < 0.05) and emotional intensity was associated with fewer CAP1 occurrences (b = -0.23, *t* = -3.17, *p* = 0.002, FDR_total_ *p* < 0.05). Multivariate LMMs confirmed a specificity between visual properties and CAP4 (b = 0.37, *t* = 2.77, *p* = 0.007), but did not demonstrate a specificity between emotional intensity and CAP1 (b = -0.09, *p* = 0.530; Table 2).

**Table 2.**
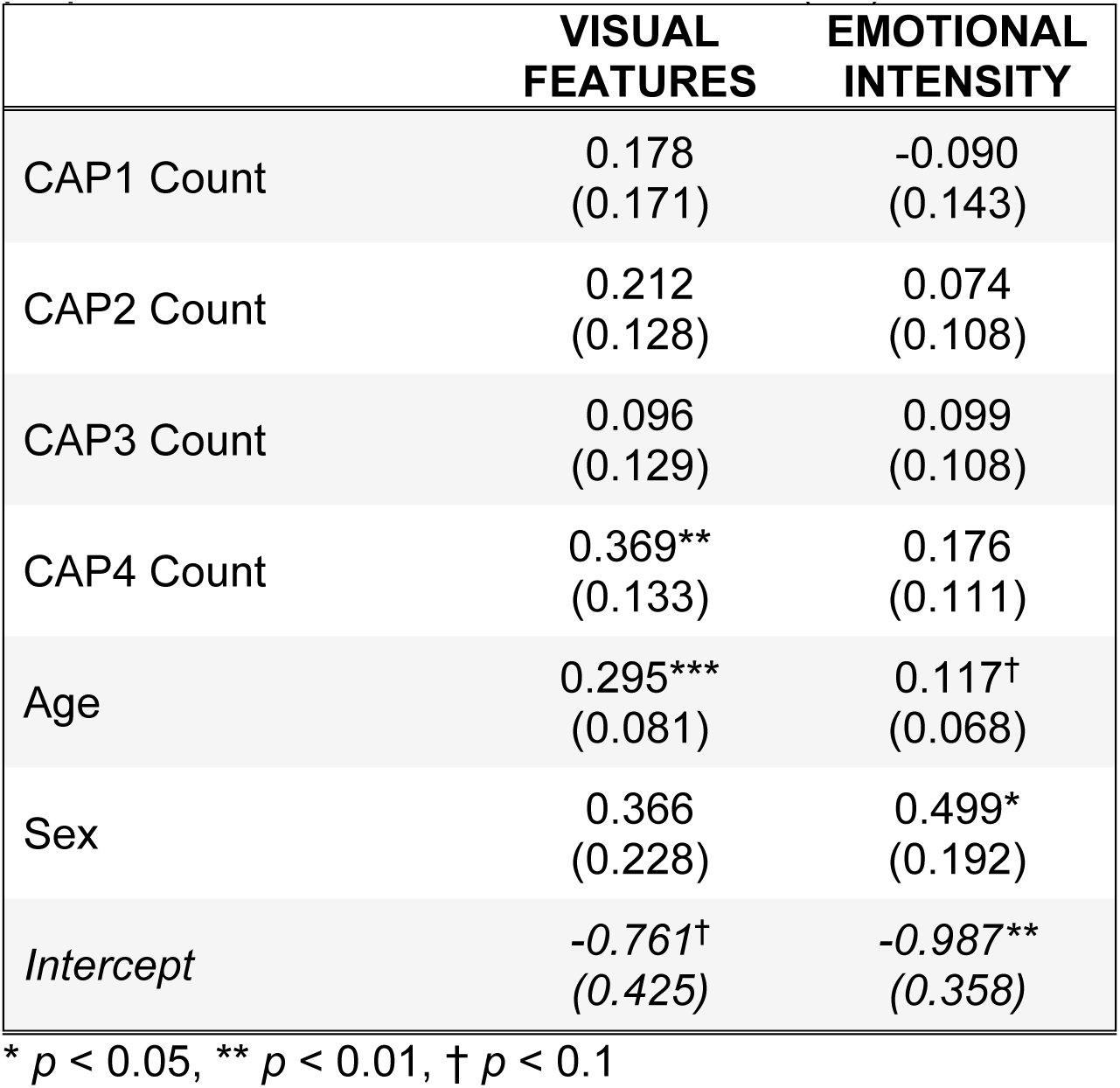
Multivariate linear mixed effects models of significant TR-IM properties for all CAPs Count. Estimates (*SE*).

### CAP Persistence and TR-IM Properties

Reliving was associated with more persistence of CAP2 (pHPC-VC; *r*_partial_ = 0.28, *p* = 0.009, FDR_reliving_ p < 0.05; Figure 3B) and emotional intensity was associated with less persistence of CAP1 (aHPC-DMN; *r*_partial =_ -0.30, *p* = 0.007, FDR_emo_ < 0.05; Figure 3D). Individual linear regressions confirmed a specificity between reliving and CAP2 persistence (b = 31.94, *t* = 2.22, *p* = 0.029). The specificity between emotional intensity and CAP1 was non-significant (b = -13.289, *t* = -1.75, *p* = 0.084), with an additional trending association between emotional intensity and CAP4 (b = 25.96, *t* = 1.82, *p* = 0.073).

**Figure 3.**
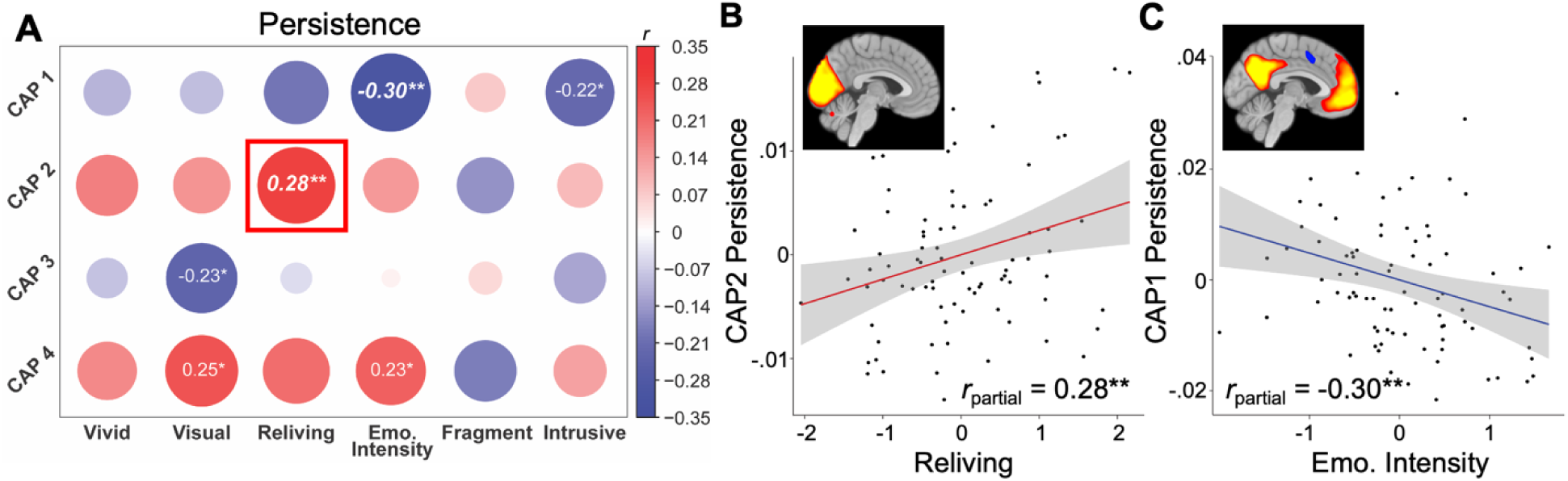
Associations between CAP Persistence and TR-IM properties. A) Partial correlations between persistence of all CAPs and TR-IM properties, controlling for sex and age, with the specific scatter plots of B) CAP2 persistence and reliving and C) CAP1 persistence and emotional intensity. Italics denote associations surviving correction for multiple comparisons. Boxes denote associations that were significant in multiple linear regression models, demonstrating specific association between that CAP and TR-IM property. * *p* < 0.05, ** *p* < 0.01.

Results from univariate LMMs confirmed these associations – reliving was associated with CAP2 persistence (b = 35.41, *t* = 2.66, *p* = 0.009) and emotional intensity was associated with CAP1 persistence (b = -18.32, *t* = -2.86, *p* = 0.005). Multivariate LMMs confirmed a specificity between reliving and CAP2 persistence (b = 33.11, *t* = 2.24, *p* = 0.028), but did not demonstrate a specificity between emotional intensity and CAP1 persistence (b = -12.19, *p* = 0.108; Table 3).

**Table 3.**
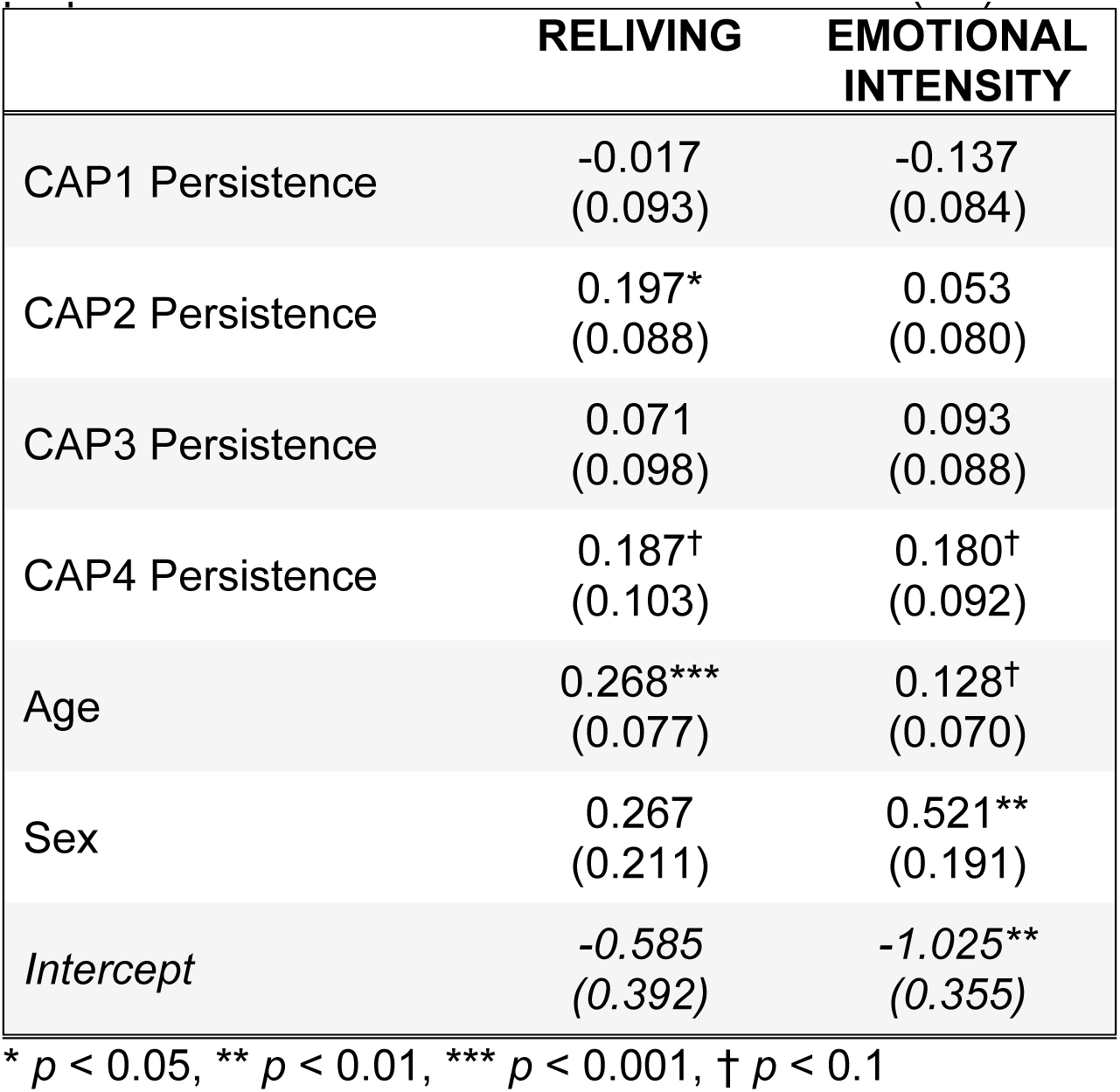
Multivariate linear mixed effects models of significant TR-IM properties for all CAPs Persistence. Estimates (*SE*).

### Associations with conventional clinical assessments

No associations were found between CAP metrics and the total number of TR-IMs during the EMA period (absolute *r*’s < 0.12, *p*’s > 0.288). Similarly, no associations were found with PTSD symptom clusters or total PTSD symptom severity (absolute *r*’s < 0.15, *p*’s > 0.166). Weak associations that did not survive correction for multiple comparisons were seen with retrospective reports of TR-IM properties on the AMQ at Visit 2 – reliving was associated with fewer occurrences of CAP1 (*r* = -0.24, *p* = 0.031) and emotional intensity was associated with less persistence of CAP1 (*r* = -0.24, *p* = 0.034). No other effects with retrospective reports of TR-IM properties were seen (*p’*s > 0.07).

## DISCUSSION

The primary aim of the present study was to utilize ecological assessments of the phenomenological properties of TR-IMs to shed further light on their underlying mechanisms, specifically with regards to HPC-cortical interactions. Consistent with our hypotheses, divergent patterns of HPC-cortical resting-state coactivation were associated with different TR-IM properties. Emotional intensity was associated with less frequent and persistent coactivation of the aHPC and DMN, while visual features were uniquely associated with more frequent coactivation of the HPC with sensory cortices and the ventral attention network. Additionally, reliving was associated with more persistent, but not frequent, co-activation of the pHPC and visual cortex. These findings align with prior work that demonstrates different HPC-cortical systems support different multidimensional features of episodic memory, such as basic sensory-perceptual versus more complex cognitive-affective details. Moreover, our findings provide novel evidence for the involvement of these different systems in the unique phenomenological properties of TR-IMs, which are core symptoms of PTSD.

To our knowledge, this is the first examination of CAPs in the context of PTSD and its core symptomatology. CAP analyses have been increasingly used in the investigation of mechanistic processes underpinning various psychiatric disorders given their sensitivity to meaningful network dynamics [67–69]. Here, the use of CAP analyses allowed investigations into the dynamic properties of these large-scale distributed networks and their reciprocity with HPC activity in a data-driven manner. The intrinsic functional architecture of the human brain is supported by evolving and dissolving “states” or patterns of coactivation that constitute canonical resting-state networks [70]. These canonical networks emerged within our data, including the DMN, D/VAN, SN, and visual network. The transient nature of these network configurations is believed to facilitate rapid and efficient information processing along the cortical hierarchy [71]. This function is of particular relevance for memory-related processes, given the widely distributed spatiotemporal networks involved in memory. It has been argued that static investigations of these networks averaged across time points result in the loss of valuable information, both at the neural and behavioral levels [60, 72, 73], thus emphasizing the importance of dynamic measures such as CAP analyses. While some studies have shown that CAPs may not represent these dynamic spatiotemporal properties of distinct network states [74], more recent work utilizing similar co-activation methodology has demonstrated meaningful temporal evolutions of network states that map onto temporally-varying behavioral processes [70]. Balancing these perspectives, we avoid the term “states” in reference to CAPs and discuss the frequency and persistence of these co-active patterns over time.

The most prominent and persistent co-activation pattern (CAP1) consisted of aHPC activation with the DMN and deactivation of attentional networks, reflecting a canonical “resting-state” pattern of activity. The integrity of the anticorrelation between attention-related networks and the DMN serves a critical role in supporting various cognitive-affective processes [75, 76], and its disruption has been linked to numerous psychiatric disorders [77–79]. The DMN has gained increasing recognition as an active player in various cognitive processes, particularly memory. Hubs of the DMN are at the center of a controlled “cortical memory retrieval network” [43] and are situated immediately downstream the HPC in a cascaded memory replay system [80]. The HPC and DMN demonstrate reciprocal interactions in the volitional retrieval of memory [23], supported by their robust intrinsic connections via the aHPC [28, 29]. The aHPC-DMN circuit in particular has been linked to the reconstructive recall of autobiographical memories, specifically the overall schematic “gist” [26, 81]. Moreover, the core DMN system responds to the affective valence, but not vividness or detail, of mentally reconstructed events [82] and both the aHPC and DMN contribute to the affective processing of past emotionally-laden experiences [83, 84]. Taken together, this aligns with our findings that link emotional intensity of TR-IMs to this aHPC-DMN CAP, suggesting this pattern of hippocampal-cortical interactions may be responsible for the affective features of autobiographical memory. A disruption in the frequency or stability of these interactions may reflect a breakdown in the intrinsic control of such affective memories, thus increasing susceptibility to spontaneous intrusions of the affective features of autobiographical memories.

Paralleling these affective properties, the sensory features of TR-IMs were associated with a pattern of co-activation across the sensory cortices and VAN/SN (CAP4). Our probes of the sensory properties of TR-IMs focused on visual features given the predominant role of mental imagery in IMs [6]. While CAP4 was marked by co-activation of the visual cortex, there were similar activations across the somatosensory and motor cortices, reflecting multimodal sensory activity in relation to the sensory (visual) properties of TR-IMs. These findings are well-aligned with prior work demonstrating an active role of the broader sensory cortex in prompted trauma-memory recall and general episodic memory replay, with activations spanning multiple sensory modalities involved in the initial processing of the stored event [85–87]. CAP4 was also characterized by co-activation of the VAN, including the dACC and AI, which comprise the highly similar SN. These networks are associated with bottom-up, sensory-driven attentional capture and are implicated in the processing of multimodal sensory stimuli [88–90]. Notably, both the sensory cortices and hubs of the VAN/SN have been theorized as neural substrates of the “sensory-representation system” in the dual-representation theory of IMs. Here, we provide critical neurobiological evidence for the co-activation of these networks in such sensory properties of TR-IMs and offer novel empirical support for this facet of the dual-representation theory.

Surprisingly, the reliving properties of TR-IMs were associated with the persistence of co-activation of the visual cortex and the pHPC (CAP2). This CAP was hypothesized to support the sensory features of TR-IMs, given the role of pHPC-VC interactions in detailed mental imagery and recall of specific sensory details of memory [26, 27, 91]. Nonetheless, visuospatial details are known to contribute to the experiences of reliving the traumatic event in the here-and-now, characteristic of severe TR-IMs and their counterpart, “flashbacks” [92]. Moreover, extant models of flashbacks and the reliving of traumatic events reliably implicate activation of the visual system [93, 94], and reconstructive recall of events, a process involving the detailed reexperiencing of individual episodes, is supported by the pHPC [21]. Notably, pHPC interactions with visual areas have been found to support the elaboration, or mental reliving, of autobiographical memory through the recovery of sensory details [95]. Therefore, the persistence of co-activation between the pHPC and VC, even at states of rest, may bias sensory-driven reconstructive recall of a traumatic event and contribute to the spontaneous reliving of TR-IMs in the “here-and-now”.

We did not find associations between HPC-cortical networks and memory vividness and fragmentation. Interactions between the pHPC and posterior midline structures have been previously implicated in the vividness of episodic memory recall and related processes of mental imagery [26, 31]. However, some data suggest the vividness of episodic memory may be mediated by cortical structures independent of the HPC, specifically the PCC/Precuneus, angular gyrus, and fusiform gyrus [96–98]. Conversely, the HPC has been more reliably implicated in the binding of disparate episodic memory details into a coherent memory representation and is thus viewed as a hub for memory fragmentation, or lack thereof [20]. Therefore, it is possible unitary HPC dysfunction may underlie fragmentation, rather than its interactions with large-scale networks. Alternatively, the role of HPC-cortical interactions in either vividness or fragmentation of memory may be task-dependent and not contingent on the intrinsic functional architecture of these networks. Therefore, future studies utilizing task-based investigations of a/pHPC-cortical network dynamics are needed to ascertain their role in vividness and fragmentation.

With this in mind, the present study has a series of limitations. Notably, fMRI analyses were constrained to the resting state, and data were collected on the order of days to weeks after the completion of the EMA surveys. While it is clear the ecological assessments of TR-IM properties yielded valuable information, as no effects were seen with retrospective recall measures, functional imaging of HPC-cortical network dynamics during actual memory retrieval or replay may yield more nuanced insights into the neural substrates of these different memory processes. Moreover, the dynamic nature of our CAP analyses may allow for identification of changes in neural “states” in response to spontaneous memories during the resting-state [72]. Indeed, there is evidence to suggest the presence of numerous ongoing cognitive processes during a “resting-state” that can be reliably detected and measured using periodic prompts [99]. Therefore, future studies examining these CAPs during either symptom provocation paradigms, prompted memory retrieval, or periodic probing for spontaneous memory emergence are warranted. Similarly, assessments of sensory-perceptual properties of TR-IMs beyond the visual system are needed, including somatosensory, auditory, and olfactory, as well as interoceptive sensations [14, 100, 101]. Additionally, our sample was predominantly female, precluding any investigations into sex differences despite known effects of sex on PTSD symptoms, sensory processing, and cognition. While we controlled for sex in our analyses, future studies matching groups by sex may yield more detailed insights into sex differences in the neurobiological substrates of TR-IMs.

Overall, our findings provide novel insights into the mechanistic processes of TR-IMs and elucidate unique neural network dynamics underpinning their phenomenological properties. The shared and unique co-activation patterns of the aHPC and pHPC lend further credence to their functional specialization with respect to large-scale neural networks and related aspects of memory. Their unique associations with different TR-IMs properties shed further light on previously observed heterogeneity in symptom-mechanism associations in trauma-related disorders. Moreover, our data demonstrate the clinical relevance of ecologically-valid assessments of the idiosyncratic manifestation of intrusion symptoms, as no symptom-mechanism associations were seen with retrospective recall measures of TR-IMs. Together, these data position dynamic HPC-cortical networks as viable intervention targets for transdiagnostic TR-IMs. Indeed, recent developments of non-invasive brain stimulation and neurofeedback have successfully targeted the identified networks [102, 103] and demonstrated its utility in the prophylactic reduction of IM intensity [16]. The incorporation of these neuromodulatory techniques targeting these networks with detailed assessments of TR-IM properties may yield individualized, mechanism-based therapies for this pervasive yet difficult to treat symptom.

## Data Availability

Data in the present study are available upon reasonable request to the authors

## ACKNOWLEDGMENTS

This work was supported by NIH awards P50-MH115874 (IMR) and R01-MH120400 (IMR).

## MATERIALS AND METHODS

### Participants

Inclusion criteria included ability to provide written informed consent, 18-65 years old, regular access to a smartphone to complete EMA surveys, and completion of 70% of daily EMA surveys. Participants were screened for TR-IM frequency and were considered eligible if they experienced 2 trauma-related intrusive memories (TR-IMs) in the past week. Exclusion criteria included left-handedness, medical conditions that would confound results, such as a seizure or other neurological disorder, history of moderate to severe traumatic brain injury, MR contraindications, including metal implants and claustrophobia, positive pregnancy test for female participants on the day of scanning. In addition, participants were excluded for current (past month) moderate-to-severe alcohol or substance use disorder, psychotic disorder, or manic or mixed mood episode. The present study utilized data from a larger study and focused only on participants with neuroimaging data.

### MRI data acquisition and preprocessing

MRI was conducted using the HCP Lifespan protocol [1]. T1-weighted 3D MPRAGE structural images were acquired using the HCP 0.8mm resolution sequence (TR/TEs: 2500/1.81/3.6/5.39/7.18; flip angle: 8 deg; FOV: 256 x 240; voxel size: 0.8mm isotropic), and eyes-open resting state T2-weighted echoplanar images were acquired using the HCP Lifespan sequence (TR/TE: 800/37 ms, in-plane resolution: 2mm; voxels: 2mm isotropic; multiband factor = 8; anterior-posterior phase encoding; one run of 976 frames, ∼13 minutes in length).

T1-weighted (T1w) images were corrected for intensity non-uniformity [2]. Brain tissue segmentation of cerebrospinal fluid (CSF), white matter (WM) and gray matter (GM) was performed on the brain-extracted T1w [3]. Brain surfaces were reconstructed using recon-all [4], and the brain mask estimated previously was refined with a custom variation of the method to reconcile ANTs-derived and FreeSurfer-derived segmentations of the cortical GM of Mindboggle [5]. Volume-based spatial normalization to MNI standard space (MNI152NLin6Asym) was performed through nonlinear registration with antsRegistration (ANTs 2.3.3), using brain-extracted versions of both T1w reference and the T1w template.

EPI images were corrected for susceptibility distortions using the fMRIPrep fieldmap-less approach [6]. Based on the estimated susceptibility distortion, a corrected EPI (echo-planar imaging) reference was calculated for a more accurate co-registration with the anatomical reference. The reference was co-registered to the T1w reference with six degrees of freedom [7]. Head motion parameters with respect to the reference (transformation matrices, and six corresponding rotation and translation parameters) were estimated before any spatiotemporal filtering [8]. EPI images were slice-time corrected [9]. The time series were resampled onto their original, native space by applying a single, composite transform to correct for head motion and susceptibility distortions. The time series were resampled into standard space, generating a preprocessed run in MNI152NLin6Asym space. First, a reference volume and its skull-stripped version were generated using a custom methodology of fMRIPrep. Automatic removal of motion artifacts using independent component analysis (ICA-AROMA) [10] was performed on the preprocessed images on MNI space time-series after removal of non-steady state volumes and spatial smoothing with an isotropic, Gaussian kernel of 6mm FWHM (full-width half-maximum). Corresponding “non-aggressively” denoised runs were produced after such smoothing [10].

### a/pHPC ROIs

a/pHPC ROIs were defined using the procedure outlined in Chen & Etkin (2013) [11]: HPC ROIs from the SPM12 Anatomy Toolbox were segmented along the anterior-posterior (Y) axis based on the following MNI coordinates: -10 to -21 (aHPC) and -32 to -43 (pHPC). These coordinates follow relevant gene-expression findings and anatomical landmarks, such as the uncal apex, while further controlling for overlap within HPC subregions and neighboring structures (i.e., amygdala).

### Co-activation Pattern Analysis

Consensus clustering was run for *k* values of 2-11 based on prior work [12, 13] and theorized limits to the number of unique brain states at rest associated with significant co-activation of the HPC. Consensus clustering was run over 20 folds, each utilizing 80% of the data, for each k individually. For each *k*, a consensus quality was computed using 1 minus the proportion of ambiguously clustered pairs (PAC) [14], with higher values reflecting more consistent clustering across folds. As this quality index increases with the number of clusters used, we subtracted a fitted exponential function from the actual data to identify the *k* that most exceeded the expected trend [15, 16]. The combination of these techniques identified *k* = 4 as the optimal number of CAP networks (Figure 1).

**Figure S1.**
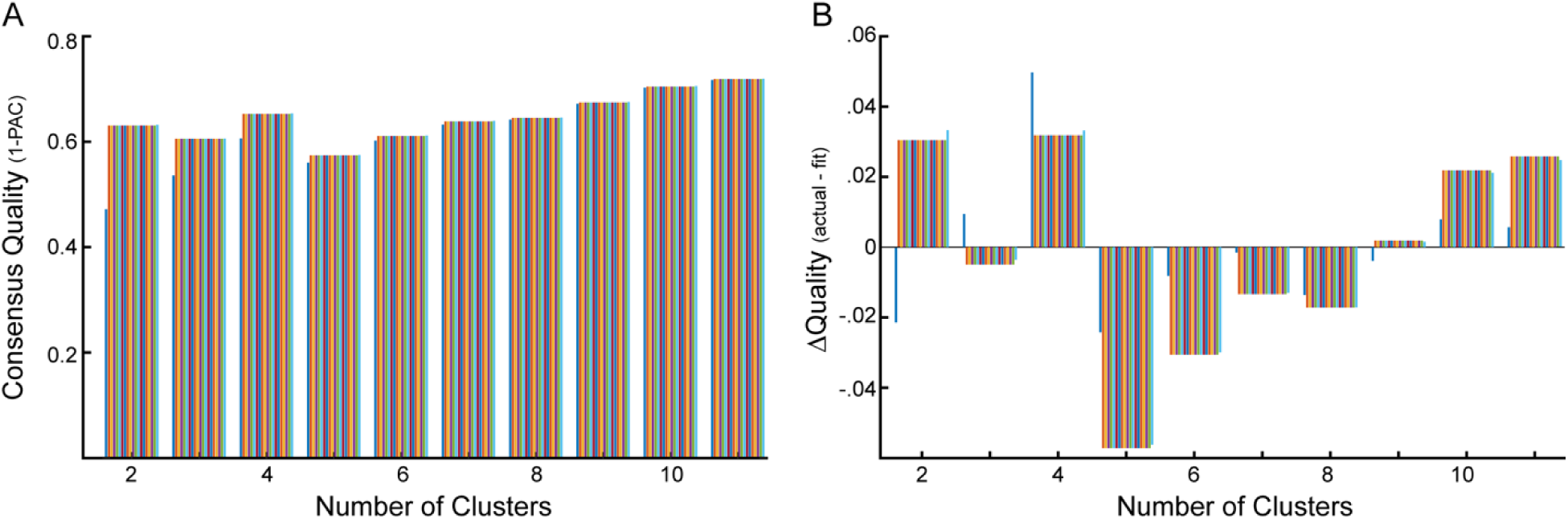
CAP consensus clustering. A) Clustering quality as measured by 1 minus the proportion of ambiguously clustered pairs (PAC) across candidate cluster numbers. *k* = 4 stands as local peak, followed by expected exponential increase with increasing number of clusters. B) Subtracting a fitted exponential function from the quality index confirms *k* = 4 as optimal. The gradient displayed for each *k* value cluster denotes different criteria for defining “ambiguous clustering”, moving along a left-right gradient from less to more strict thresholding.

## RESULTS

### Fractional count

The average number of volumes exhibiting supra-threshold activation of the a/pHPC across participants was 264.2 volumes (SD = 11.8). Effects using fractional, instead of raw, CAP counts demonstrated virtually identical results. Visual properties were associated with more occurrences of CAP4 (*r*_partial_ = 0.29, *p* = 0.007), and emotional intensity was associated with fewer occurrences of CAP1 ((*r*_partial_ = -0.34, *p* = 0.002).

